# Contribution of Schools to Covid-19 Pandemic: Evidence from Czechia

**DOI:** 10.1101/2021.09.28.21264244

**Authors:** Cyril Brom, Jakub Drbohlav, Martin Šmíd, Milan Zajíček

## Abstract

**Purpose:** It is unclear how much opening of schools during Covid-19 pandemic contributes to new SARS-CoV-2 infections among children. We investigated the impact of school opening with various mitigation measures (masks, rotations, mass testing) on growth rate of new cases in child cohorts ranging from kindergartens to upper secondary in Czechia, a country heavily hit by Covid-19, since April 2020 to June 2021.

**Methods:** Our primary method is comparison of the reported infections in age cohorts corresponding to school grades undergoing different regimes. When there is no opportunity for such a comparison, we estimate corresponding coefficients from a regression model. In both the cases, we assume that district-level infections in particular cohorts depend on the school attendance and the external environment in dependence on the current overall risk contact reduction.

**Results:** The estimates of in-cohort growth rates were significantly higher for normally opened schools compared to closed schools. When prevalence is comparable in the cohorts and general population, and no further measures are applied, the in-cohort growth reduction for closed kindergartens is 29% (SE=11%); primary: 19% (7%); lower secondary: 39% (6%); upper secondary: 47% (6%). For secondary education, mitigation measures reduce school-related growth 2-6 times.

**Conclusion:** Considering more infectious SARS-CoV-2 variants and the ‘long covid’ risk, mitigation measures in schools, especially in secondary levels, should be implemented for the next school year. Some infections, however, are inevitable, even in kindergartens (where mitigation measures are difficult to implement) and primary schools (where they may not work due to low adherence).

## 1 Introduction

Closing schools for in-person education during the Covid-19 pandemics presents a controversial issue. During the first half of 2020, closing schools appeared to be one of the most effective intervention to mitigate the Covid-19 spread [1–3]; however, closed schools negatively impact children [4–7]. Consequently, governments tended to open schools for the school year 2020/21, but with various safety measures; mask wearing and mass screening among others. Altogether, most of these measures appeared to reduce the risk of spreading the disease in schools [8–10]. However, there is still uncertainty how much open schools contribute to the number of child infections; especially as concerns younger cohorts, including kindergartens wherein mitigation measures are difficult to implement. Children may be less susceptible to SARS-CoV-2 infection [11, 12] (but see also [6]), but outbreaks occur even in preschool settings [13] and number of school cases tend to increase when incidence in general population increases [14–16].

Czechia was among countries worst hit by the pandemic, with 14-days incidence per 100k inhabitants >500 for most of the October 2020 – April 2021 period [17] (Figure S3). For weeks, educational institutions were closed for most children, but remained open for some; such as open for kindergartens but closed for all other educational levels from October-14-2020 to November-16-2020, or open for Grade 1-2 but closed for Grade 3-5 of the primary level from January-4-2021 to February-26-2021. Masks were required since September-17-2020 among staff and pupils in all educational levels except of kindergartens and regular mass screening of school children and staff started on April-12-2021. This created several unique “natural experiment” settings enabling to compare numbers of infections among similarly-aged cohorts that remained home versus those attending schools in person: with different mitigation measures in place.

Here, we use Czech nationwide data since April-6-2020 to July-4-2021 and a simple epidemiological-like model to infer increases in age-specific growth rate of incidence caused by in-person schooling (increases on the top of the baseline, which is distance education). In the model, the infections in the child cohorts depend on the overall magnitude of epidemic, its magnitude within the district and within the corresponding cohort, the latter in dependence on the current school opening regime. Hence, the model reflects the dependence of the effect not only on the regime introduced, but also on prevalence within the involved cohorts. We cluster children into four cohorts: kindergartens, primary schools (Grade 1-5) and lower (Grade 6-9) and upper (Grade 10-13) secondary schools.

## 2 Methods

### 2.1 Data sources

#### 2.1.1 Age and District Specific Incidence

To get the weekly numbers of reported infections for individual age cohorts, we used the national anonymized list of positively tested individuals [17] including, among other things, the date of reporting the infection and the age. There are surely unreported infections; however, as we discuss in Suppl. Mat. Section SF, our results are quite robust in this respect as we primarily examine growth rates rather than absolute numbers.

We use data since April-6-2020 (week 15/2020), as we regard prior data noisy, unreliable and suffering from small sample properties (the first case in Czechia was reported March-1-2020). The data series ends on July-4-2021 (week 26/2020). As we do not use data from weeks 27-35/2020 of summer vacation and those from the one-week winter vacation (53/20), this means 55 weeks. There are 77 districts, so we have 4235 observations, from which, however, some additional ones were excluded (see Section SB for details).

#### 2.1.2 School Opening Regimes

In addition to the complete closure, we distinguish three main regimes in which schools were opened:

- without masks wearing,
- with wearing masks for both teachers and pupils,
- with masks and a weekly rotation regime (entire class in school one week, at home the other week).

For each regime, we distinguish whether or not regular weekly antigen testing took place at schools. Which regimes took place at specific school levels is listed in Table S3.

#### 2.1.3 Contact Restriction

To control for external (out of school) influence on the infections of children, we use the estimates of overall risk contact restrictions [18] from a longitudinal covid-related study, surveying bi-weekly a representative panel of 2-3 thousand Czech citizens. This study also yields data on the weekly number of risk contacts.

### 2.2 Statistical Analysis

In Czechia, numbers of children attending particular schools levels correspond well with particular age cohorts, because the entry time to the primary school is given by law and attending school is compulsory (including the final year in kindergartens). For instance, lower primary (Grade 1-5) includes 6-11-year-olds. Seven-to-ten-year-olds are almost exclusively primary school pupils; whereas, 6-year-olds attend either first grade or the final kindergarten year, and most 11-year-olds attend either primary (Grade 5) or lower secondary (Grade 6) school. See Section SB for details.

The primary method we use to estimate the influence of individual school opening regimes is the comparison of close age cohorts for which the school regime differs by means of subtracting the observations (here infection numbers) corresponding to the same time and the same district. We identified four opportunities for such a comparison.

1. Weeks 43–46 in 2020 when kindergartens remained open while the primary school closed.
2. Weeks 1–8 in 2021 when only Grades 1–2, but not Grades 3–5, from the primary level were opened.
3. Weeks 49–51 in 2020, when Grade 9 (final of the lower secondary level) opened fully but Grades 6-8 opened with rotas.
4. Weeks 49–51 in 2020, when Grade 13 (final of the upper secondary) opened, but Grades 10-12 remained closed.

The details on the sub-cohort comparisons are summarized in Table S4. For the values of the regime dummies, see Section SB.

For the parameters which cannot be estimated this way (i.e., two close age sub-cohorts did not undergo a different regime) we used a secondary approach: the direct estimation from a regression model.

In both the methods, we examine the number of reported infections in the examined age cohort for each district and week. We assume that this number linearly depends on the previous-week numbers of infections – the overall number, the number within own district and the number within own district and own cohort. In all the three cases, these numbers are adjusted by the contact restriction reported two weeks earlier and the previous-week rate of infectiousness. Additionally, the infections in the cohort within the district depends on the influence of schools, being equal to the previous-week infections in this cohort, multiplied by the sum of dummy variables corresponding to six considered opening regimes – once the regime is applied, the variable is one while the remaining dummy variables are zero. When schools (for this cohort) are closed, all the dummies are zero. See Section SA for details and for justification of this general model.

When comparing two cohorts, we assume both to follow their own version of the linear model described above (see (4) in Section SA). The advantage here is that the regime covariates which do not differ between cohorts, cancel out, so our estimate is independent on these parameters. Moreover, even though we do not explicitly assume an identical impact of the external environment on both the sub-cohorts, these impacts are likely to be similar for the cohorts, so they more or less compensate by the subtractions, disturbing less the impact of the regime terms. Note that, once total closure is compared to some regime, the significant results of this estimation can be interpreted as a rejection of the hypothesis that the corresponding regime has no influence on infections.

The direct estimation of the linear model, on the other hand, is less reliable, as it is more dependent on the choice of the general model; however, for the coefficients which cannot be obtained by cohort comparison, it is the only choice.

Another advantage of the direct estimation is that it can be used to construct a global model of the infection in the examined cohort, distinguishing the impact of schools and that of the external environment. This can be done in the language of prevalence:

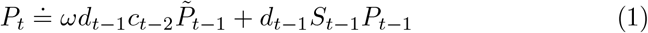

where *P*_*t*_ denotes *prevalence* within a cohort, i.e. the ratio of weekly new cases and the cohort size, and 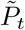 denotes the overall prevalence. The equation above may be equivalently expressed by means of growth *ρ*_*t*_ of the prevalence within the cohort:

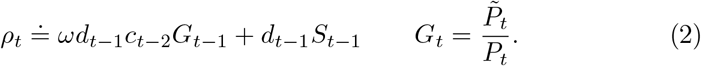

Here, *d*_*t*_ is the infectiousness in time t, *c*_*t*_ is the contact restriction (see Section SD), *ω* is an estimated constant and

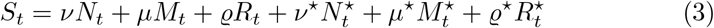

is the influence coefficient of schools, where:

- coefficient *ν* and dummy *N* correspond to the opening without masks,
- *µ* and *M* to opening with masks,
- *ρ* and *R* to rotation regime,
- and star indicates regular antigen testing at schools.

Importantly, because the model is linear, coefficients *ν*,…, *ρ*^⋆^ are mutually comparable; when *ν* is twice as much as *µ* for instance, this means that the contribution of opening without masks to the prevalence *P*_*t*_ or its growth *ρ*_*t*_, respectively, is twice as much than that of opening with masks. Moreover, these coefficients are comparable with the “external influence” coefficient *ω* provided *c*_*t*_ = 1 and 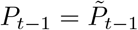, meaning once there is no contact restriction, the prevalence is the same in the cohort and generally, and, for instance, *ω* is equal to *ν*, then closing schools would reduce the cohort’s prevalence/its growth by half, or, in the situation of closed schools, opening without masks would increase the cohort’s prevalence/growth by hundred percent.

## 3 Results

We estimated the “school regime” coefficients *ν*, …, *ρ*^⋆^ by two variants of cohort comparison (if applicable) and two variants of direct estimation (see Section SA for details); consequently, we made meta-estimates out of all the available estimates. The meta-estimates are computed as weighted averages of the direct estimates and available comparison ones with strong preference of the latter (see Section SC for details). To distinguish the quality of the meta-estimates, we have split them into the following four categories.

● At least one source estimate is a comparison of a regime and the closure, where both the cohorts correspond to the same school level and are “interior”, i.e. 7-year-olds (Grade 1/2 open) vs. 9-year-olds (Grade 3/4 closed).
◕ At least one source estimate is a comparison of a regime and the closure such that either the comparison includes cohorts from different school levels (i.e. 5-year-olds [kindergarten open] vs. 7-year-olds [Grade1/2 closed]) or a “boundary” cohort (i.e. only the last grade of some school level was opened while the rest was closed). In the latter case, because no age cohort existed that would include only individuals from the open grade, halves of the boundary cohorts had to be used.
◑ At least one source estimate is a cohort comparison, but of two different regimes (e.g. masks and rotations).
◔ The meta-estimates based only on direct estimation from the regression model.

Clearly, the better category, the more reliable the estimate. The resulting estimates are listed in Table 1 and Figure 1. Missing estimates refer to situations when schools were never opened in corresponding regimes. Apparently, upper secondary school coefficients are larger than lower secondary school coefficients, which are larger than primary school and kindergarten coefficients. For secondary education levels, mitigation measures reduces the coefficients notably; whereas, for primary schools, they remain similarly low no matter the regime. (For kindergartens, no mitigation measures were implemented: they were either open or closed.)

**Table 1.**
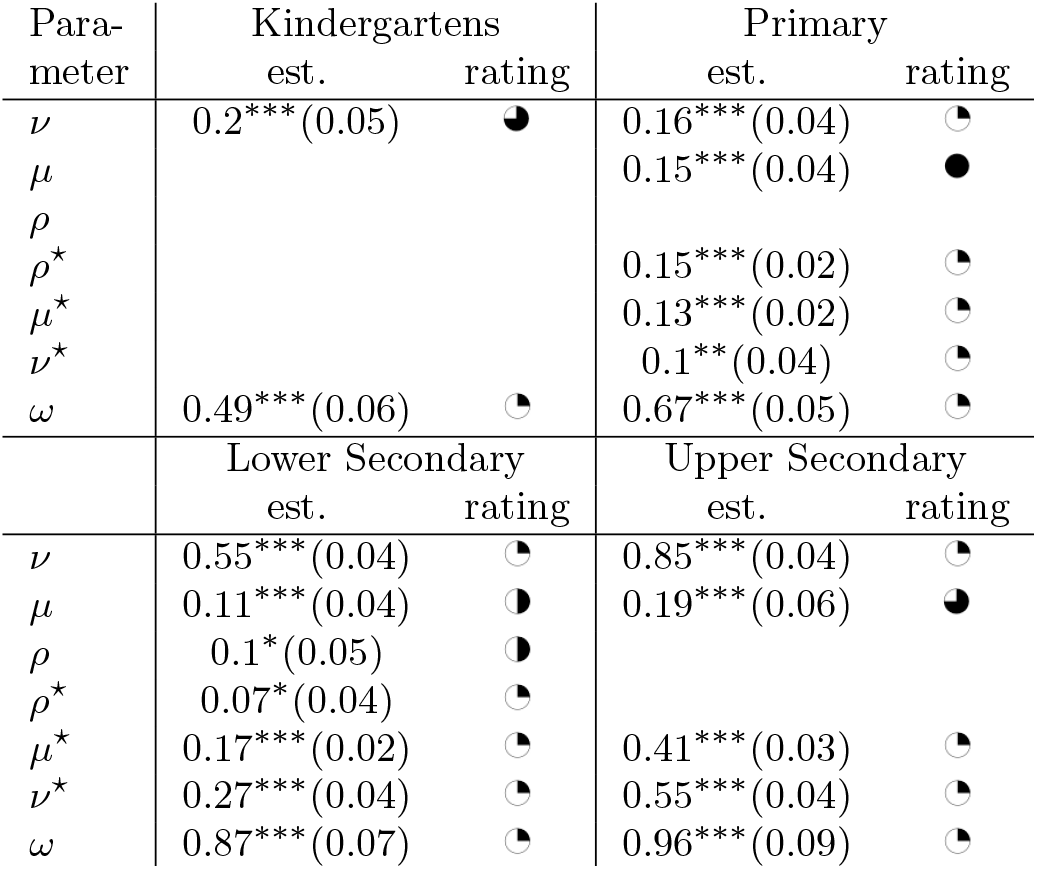
Meta-estimates of regime coefficients (SE in parentheses). * *p <* 0.10, ** *p <* 0.05, *** *p <* 0.01 of the two-sided hypothesis.

**Fig. 1.**
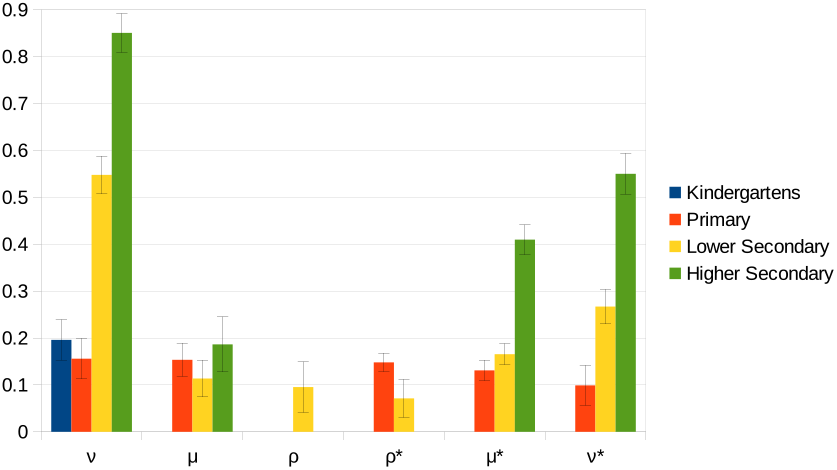
Meta-estimates of regime coefficients

Further, for each school level and each regime, we evaluated the mean changes to the growth in the corresponding cohort associated with imposing the regime. In particular, we computed the hypothetical, mean relative increase *r* of the growth after opening closed schools in that regime, and a hypothetical, mean relative decrease *e* of the growth after imposing the regime into a school opened without any mitigation measure. These quantities can be computed for various *c*_*t*_ (contact restriction) and *G*_*t*_ (prevalence ratio, Eq. 2); here, for simplicity, we assumed identical previous prevalence in and outside the cohort (*G*_*t*_ = 1) and no contact restriction (*c*_*t*_ = 1), in which case 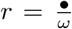 and 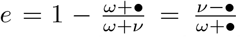 where • denotes the corresponding regime coefficient. The results are listed in Table 2. Again, we see that the impact of open schools is larger for older students than for younger ones, but mitigation measures reduce school-related growth substantially for the latter group. At the same time, school-related growth in kindergartens appears to be larger than growth in primary schools, but the difference is not significant.

**Table 2.**
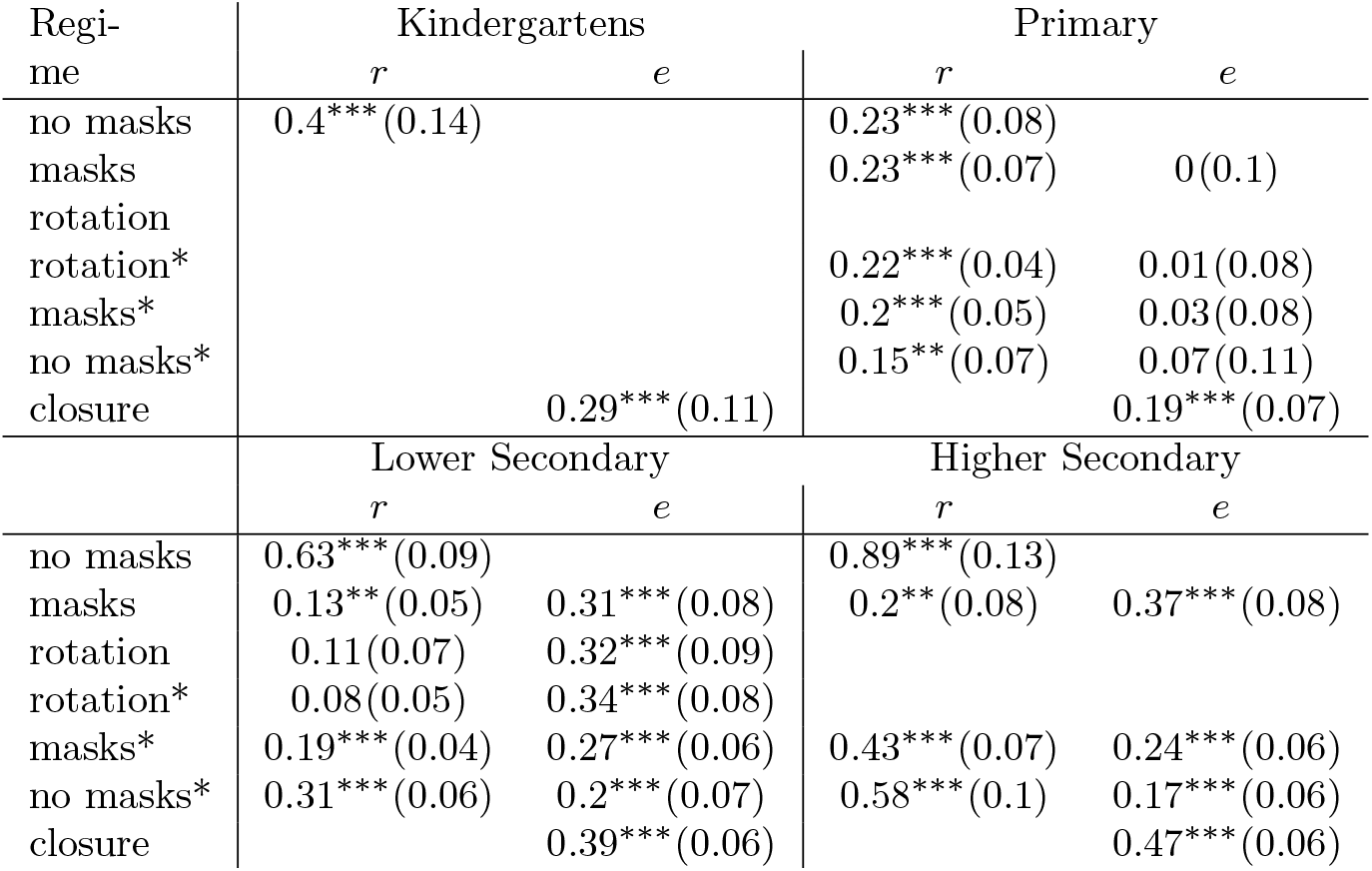
Effects of individual regimes. *r* – relative increase of prevalence within the cohort given no contact restriction, the same cohort and general prevalence, and schools closed, *e* – relative decrease of prevalence within the cohort given no contact restriction, the same cohort and general prevalence, and schools open. Upper estimate of standard errors in parenthesis. * *p <* 0.10, ** *p <* 0.05, *** *p <* 0.01 of the two-sided hypothesis.

Finally, for each cohort and time *t*, we evaluated *ρ*_*t*_ serving as a prediction of the actual growth rate 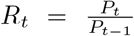 computed by means of the meta-estimates (i.e., those in Table 1). The results are depicted on Figure 2 together with the decomposition of the (predicted) growth into the parts caused by individual regimes (i.e., the non-red parts on Figure 2) and the external, out-of-school environment (i.e., the red part). Unlike Table 2, Figure 2 depicts the situation after accounting for changing contact restrictions in time and differing prevalence in individual child cohorts and general population. Hence, as concerns description of the situation in Czechia, Figure 2 charts more accurate picture than Table 2 alone. For instance, it can be noticed that even after accounting for contact restrictions and differing prevalence, contribution of open schools to growth in child cohorts was substantial. However, it was almost always lower compered to growth due to external, out-of-school environment.^1^

**Fig. 2.**
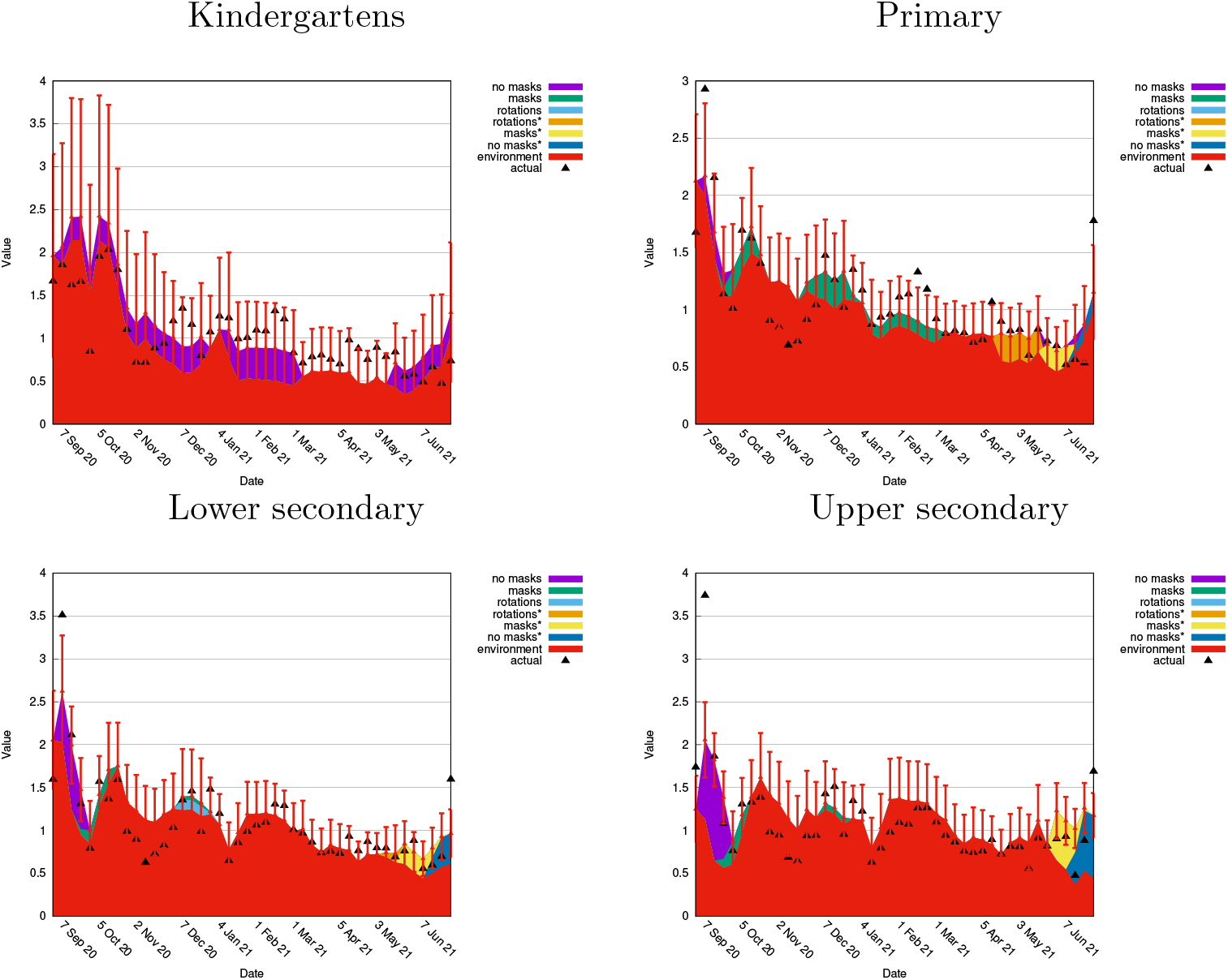
Growth 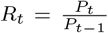, its one-week prediction *ρ*_*t*_ and its decomposition. Errorbars: 95% prediction CI. Note that when the graph area is just red in a specific time, this typically means schools were closed that time.

## 4 Discussion

The results suggest that opening any school level, including kindergartens, influenced the infections within corresponding student cohorts in Czechia. Yet children from all cohorts appeared to be infected more often outside schools then in relation to open schools. This picture agrees with the emerging view that Covid-19 infections in schools do occur, they can occur frequently when the background incidence in population is high, but schools are unlikely the key driver of the epidemic [6, 8, 10, 14, 16].

The results indicate that when lower and upper secondary schools are open, mitigation measures in these schools reduce the part of the growth which is school-related roughly 2-6 times, the effect of most effective measures not being much different from total closure (total closure would mean zero school-related growth). Such a reduction, however, is not apparent in primary schools (possibly because school-related growth is generally low therein). On the one hand, our results are not robust enough to pin down effectiveness of individual measures. Plus, our estimates for schools run without any mitigation measure are least robust. On the other hand, our estimated for schools run with face masks are more robust. Hence, altogether, these results support another emerging idea: that schools run with various mitigating measures, including face masks, have tens of percent lower risk of infection for students [8, 9, 19], but some of these effects may be weaker for primary schools [9].

Relatively low school-related growth for primary schools in Czechia agrees with other literature [8, 20]. A new, surprising finding concerns kindergartens: the school-related growth rate does not appear to be particularly low therein. This supports another emerging idea that the difference in susceptibility/infectiousness between young children and adults may be lower than thought at the beginning of the pandemic (see [6]). Plus, biological susceptibility/infectiousness can be obscured in kindergartens by these children being less able to adhere to hygienic norms and to keep distance among themselves and teachers.

The straightforward implication from this study is that when schools, especially upper grades, are open for in-person education, running them with a combination of mitigation measures is helpful. However, it is important not to forget two things. First, success of masks wearing and testing depends on adherence. Second, effectiveness of rotations is also based on restrictions outside schools (which were notable in Czechia when rotations were implemented). Reducing the risk of Covid-19 spread in schools is especially important in the light of more infectious variants of SARS-CoV-2 and the majority of children being unvaccinated. Mortality and hospitalization among young people is low [6], but ‘long covid’ could burden a notable proportion of infected children [21, 22] (but see also [23]). Hence, the case of kindergartens and primary schools is a challenging one because mitigation measures are difficult to implement effectively therein.

The strength of the study lies in that it is based on over-year-long data series from a national database and a longitudinal study concerning behavior of Czech citizens during the pandemic. Plus, some of the robust estimates were drawn because of the unique Czech situation: some school grades were opened whereas others remained closed when general incidence was very high. The strengths also include robustness of the model with respect to various ad hoc specified external parameters (apart from infectiousness; see Section SE for the respective sensitivity analysis). Our results are relatively robust with respect to the fact that some cases remain unreported (the ascertainment rate is non-unit). The main reason for this robustness is that we primarily study relative growth which is more or less independent of the ascertainment rate. Section SF shows that the fact that some cases are not reported either does not disturb the analysis at all or our estimation procedures may be modified to accommodate this fact. For instance, under the natural assumption that school regimes including regular testing increase the ascertainment rate, our results do not change substantially (Section SF).

As in any “natural experiment” study, the general limitation is that the comparison cohorts are not truly randomized. Hence, infection numbers in these cohorts may be differentially influenced by uncontrolled factors. Evidence from other countries could strengthen the reliability of our results. Also, our estimates (Tables 1, 2 and Figure 1) apply to situation without contact restrictions and the same prevalence in and outside the examined cohort; so rather than their value, their ordering and ratios should be interpreted. Nevertheless, our model itself is general and when using actual prevalences and contact restriction, it can predict growth as we did on Figure 2. Magnitudes of these particularized predictions (i.e., on Figure 2) do not differ much from the crude estimates (i.e., those in Tables 1, 2 and on Figure 1). This is because for most of the examined period in Czechia, the higher overall prevalence in the population than in the examined cohorts (except for upper secondary level) had the tendency to counterbalance effects of reduced contacts (Figures S4, S6).

In conclusion, our findings indicate that school opening have a notable effect on school-related growth rate of infections, but it can be substantially reduced by means of mitigation measures, especially in secondary education levels. The challenging question remains how to increase safety in kindergartens and primary schools wherein mitigation measures appear to be difficult to implement effectively.

## Data Availability

All data are available on https://github.com/cyberklezmer/schoolpaper.

## 5 Declarations

### 5.3 Availability of data and code

All data are available on https://github.com/cyberklezmer/schoolpaper.

### 5.4 Ethics approval

Ethics approval was waived because the study works only with anonymized, national open data.

## Acknowledgment

The authors are grateful to Ján Palguta and Arnošt Komárek for valuable comments and suggestions.

## Supplementary material

### A Methods

The primary method we estimate the influence of attending schools is the comparison of similar age cohorts for which the school regime differs. For the parameters which cannot be estimated this way we use a secondary method: the direct estimation from a regression model.

In both the methods, we primarily examine the number of reported infections *X*_*i,t*_ in the corresponding (single- or multiple-year) age cohort for each district *i* at time *t* (time granularity is one week). Generally, we assume that this number depends on the previous-week number of infections, the contact restriction reported two weeks earlier, and the previous-week rate of infectiousness. In addition to the total number of infections in population, we take into account the number of the infections in the same district, and the number of the infections in the same cohort within the district. See Section D for the justification of this general model. In addition, we extend this general model by explicitly modeling the influence of school opening regime on the infection number in the cohort.

In particular, we assume

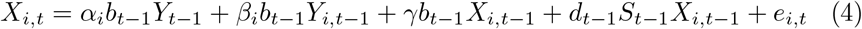

for each district *i* and time *t*. Here,

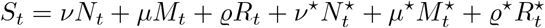

is the school influence term (also defined by (3)), *Y*_*t*_ is the number of new infections within the entire population at *t, Y*_*i,t*_ is the number of new infections within the *i*-th district at *t, N*_*t*_, *M*_*t*_, *R*_*t*_ evaluate the degree of school opening without masks, with masks, with weekly rotations (with masks on), respectively (zero means all classes closed, one means all classes open), 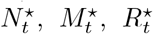 stand for the same regimes with the addition that students are regularly tested at schools by antigen tests, *e*_*i,t*_ is the error term, and *α*_1_, …, *α*_*n*_, *β*_1_, …, *β*_*n*_, *γ, ν*, …, *ϱ*^⋆^ are estimated parameters. Finally, *b*_*t*_ = *d*_*t*_*c*_*t*−1_ is the restricted infectiousness where *d*_*t*_ is the rate of infectiousness and *c*_*t*_ is the contact restriction at *t*, see Section D for details.

When comparing two cohorts, we assume them both to follow their own version of (4) either with the same or with different school regime coefficients *ν, µ*, …, *ϱ*^⋆^. We now describe two ways of comparison used in this work.

#### 1 Comparison of two cohorts with different regime parameters (later abbreviated as C)

Here we consider two sub-cohorts of a larger cohort, the *j*-th sub-cohort following its own version of (4):

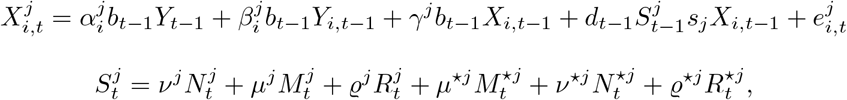

where *s*_*j*_ is the relative size of the *j*-th sub-cohort with respect to whole cohort. By subtracting, we get

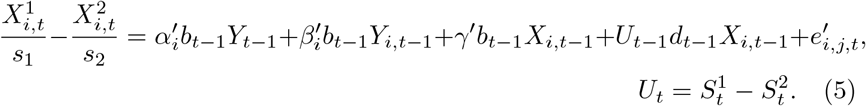

where *α*′, *β*′, *γ*′ are estimated coefficients. The advantage here is that the regime covariates and coefficients which do not differ cancel out, so our estimate is independent both of these parameters and on the choice of corresponding regime covariates, which are set subjectively sometimes, for instance when the only a subset of classes is open. Moreover, even though we do not explicitly assume an identical impact of the external environment on both the cohorts, these impacts are likely to be are similar, so they likely more or less compensate by the subtractions, disturbing less the impact of the regime terms. Clearly, for any pair of the regime coefficients out of (*ν*^1^, *ν*^2^), …, (*ϱ*^⋆1^, *ϱ*^⋆2^) to be estimable, the corresponding covariate pair (*N*^1^, *N*^2^), …, (*R*^⋆1^, *R*^⋆2^) has to differ sufficiently. This was the case, for instance, when first two grades of primary schools were opened with masks while the rest were closed 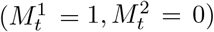 during some period and, outside this period, the whole primary schools were either open 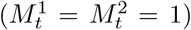 or closed 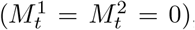, which is enough for *µ*^1^ and *µ*^2^ to be estimable (from 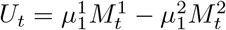, note that the rank of the corresponding covariate sub-matrix is full).^2^ Two coefficients of two different regimes can be estimated that way: when, for instance, the last grades of lower secondary were open while the rest went to rotations during a certain period, we had 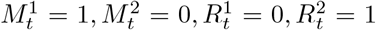 during the period; outside the period, we had 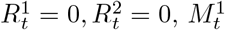 being either 0 or 1, 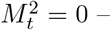 – clearly the rank of the covariate sub-matrix is full.

#### 2 Comparison of two cohorts with common regime parameters (CC)

If we decrease the generality of (5) by assuming that *ν*^1^ = *ν*^2^ = *ν*, …, *ϱ*^⋆1^ = *ϱ*^⋆2^ = *ϱ*^⋆^, we get

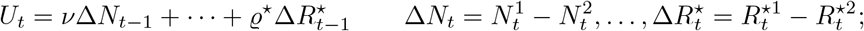

Here, the requirements for variability of covariates are less strict than above. For instance, in the case of primary schools mentioned above, it suffices that 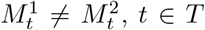 for some period *T* and 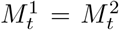, perhaps equal to zero, *t* ∉ *T*. Note that, contrary to Estimation procedure C, the estimation does not depend on values of *M* when they are equal, which may be seen as an advantage of Estimation procedure CC. On the other hand, once different regimes are applied for the two sub-cohorts, as in the example above with last grades of lower secondary schools open, and the sub-cohorts are treated equally otherwise, we get 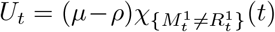 where *χ* is a characteristic function, i.e. only a difference of *µ − ρ* is estimable while the individual parameters are not identifiable. For the C procedure, things are other way around: both the regime coefficients (i.e., *µ, ρ*) may be estimated; however, observations during which the regimes did not differ have to be used.^3^

##### Other approaches

Unfortunately, the majority of the regime coefficients cannot be estimated by means of cohort comparison as no situation appeared in which the corresponding regimes differed for comparable cohorts. Thus, we have to resort to their estimation directly from the model (4) in all its generality or using its simplified version with the homogeneous impact of districts.

#### 3 Direct estimation from (4) for the whole cohort (D)

Here, all the parameters are estimated directly from (4); this, however, brings more dependence on the model, namely its part concerning external influence.

#### 4 Estimation from a homogeneous version of (4) (DH)

To get global results rather than the district-level ones, we may decrease the generality of (4) and assume a homogeneous model:

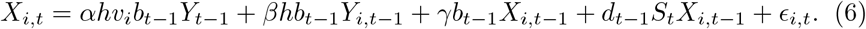

Here, *h* is the relative size of the examined cohort with respect to the rest of the population and *v*_*i*_ is the relative size of the *i*-th district’s population. By summing (6) for all *i*, we get

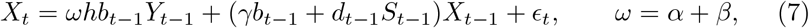

where *X*_*t*_ is number the overall cohort infections, which may be alternatively interpreted as (1) (to see it, divide (7) by the cohort size).

Finally, if we divide (1) by cohort prevalence *P*_*t*−1_, we get (2) from which we can distinguish the part of the growth caused by school opening (quantified by the second summand) from that originating outside schools (first summand). In the special case that no contact restriction is applied (*b*_*t*−1_ = *d*_*t*−1_) and that the incidence in the cohort is the same in as that within the whole population 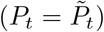, we get

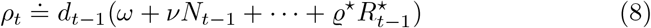

i.e. the regime coefficients themselves together with the covariates may serve as weights.

### B Data

In Czechia, numbers of children attending particular schools levels correspond well with particular age cohorts. Each September first, children who completed their sixth year are obliged to enter the first grade of a primary school. When the parents wish, a child who is six as late as by the end of the year can enter the first class, too, some children, on the other hand, start their school later for various, mostly developmental, reasons. If we neglect these minor exceptions, we can conclude that the first year pupils are six or seven years of age. Consequently, the vast majority of the primary level (Grade 1-5) pupils belong to the age cohort 6-11; the vast majority of the lower secondary school students (Grade 6-9 in Czechia) belong to the cohort 11-15; and the upper secondary school students (typically Grades 10-13 in Czechia) belong to the cohort 15-19. As for the kindergartens, where the usual lowest age for admission is three, the vast majority of children attending kindergartens falls into the cohort 4-6. The pre-school year (Grade 0) is compulsory. Younger children’s attendance in pre-school institutions is optional, but the majority of children do attend them.

For simplicity we assume that the frontier one-year cohorts split by half between the competing school categories, so the infections occurring in the cohort split by two between each category. Consequently, the number of cases by pre-school children over week *t* in the *i*-th district will be 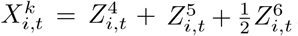, the number by the primary level 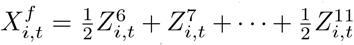, by the lower secondary level 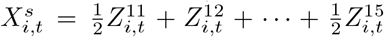 and that by the upper secondary schools 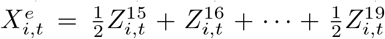,where 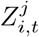 is the number of cases by the *j*-year old individuals in the *i*-th district. We take the values 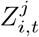 from the anonymized person-level list of reported infections [17] including, among other things, the date of reporting the infection and the age; the population and district-level series *Y*_*t*_ and *Y*_*i,t*_ are computed by their aggregation. Figure 3 displays prevalence in child cohorts and overall prevalence, Figure 4 the ratio of the child cohort prevalence to the overall one (the ratio equals to 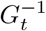) (see 2), finally Figure 6 shows fraction of cases in the child cohorts in the overall case number.

**Fig. 3.**
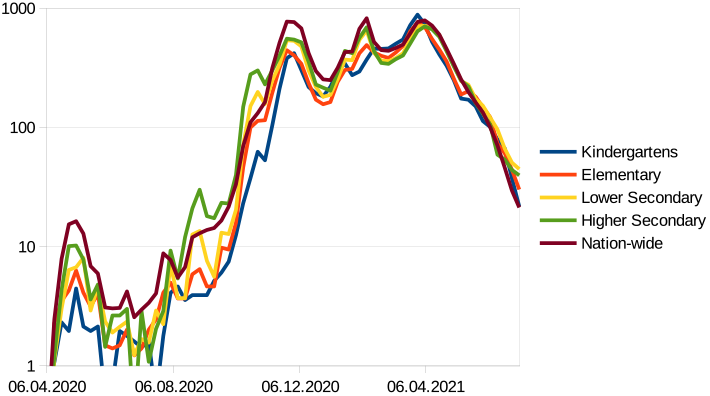
Prevalence in individual cohorts (weekly new cases per 100,000)

**Fig. 4.**
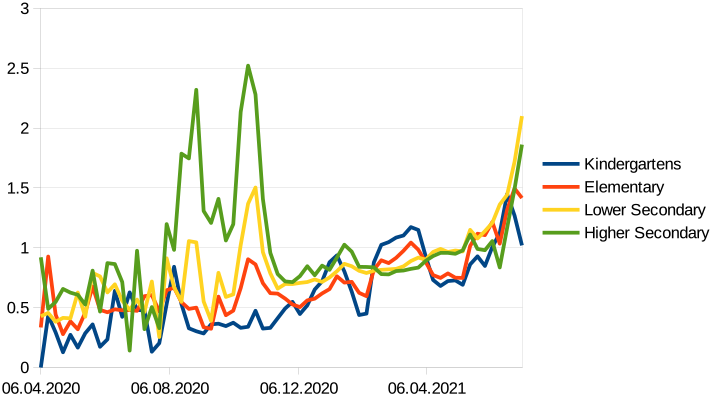
Prevalence (weekly new cases per 100,000) in child cohorts relative to overall prevalence

**Fig. 5.**
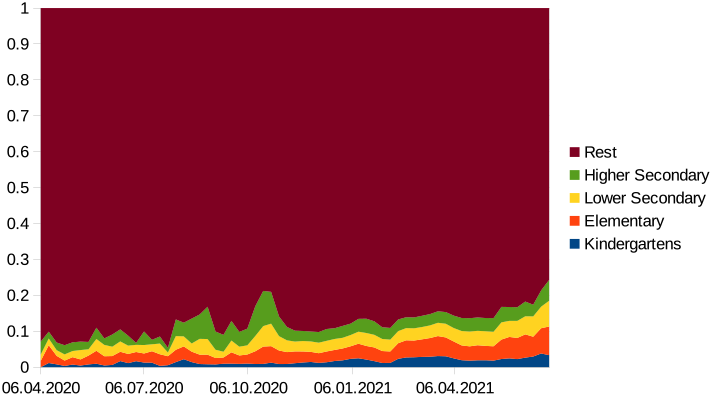
Fraction of childrens’s infections within weekly new cases

**Fig. 6.**
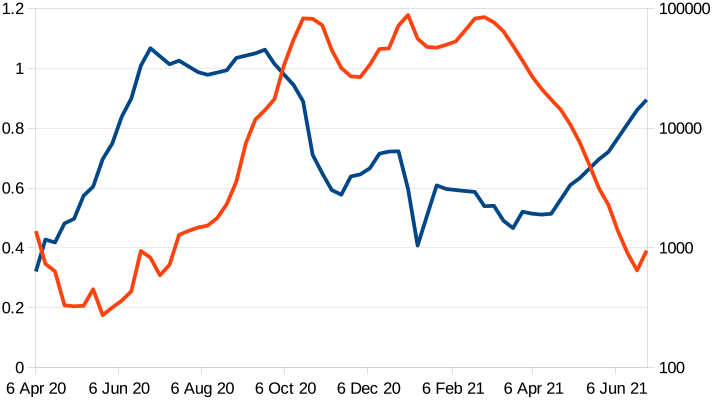
Blue: *c*_*t*−2_ – contact restriction two weeks earlier. Red: *Y*_*t*_ – weekly cases

Even though the first cases in Czechia appeared in the beginning of March, 2020, we do not take data until April-05-2020 into account, as we regard them as noisy, unreliable and suffering from small sample properties. The last values used for the estimation correspond to the week staring on June-28-2021.

The regime covariates have been determined separately for each cohort and each of the Estimation procedures C, CC, D and DH, and according to measures schedule, listed in Table 3. The values of *N*, …, *R*, which correspond to the full cohorts, were set to 1 if the corresponding regime applied fully, and to 0 if it was not applied at all in the given week. Some values were set to fractional values; some weeks were excluded from the individual estimations, especially in the cases when the fractional value could not be determined objectively. We also excluded observations corresponding to vacations (9 weeks of summer vacation in 2020 and one week of winter vacation in 2020), holidays and the weeks with no more than two possible school days, as our study concerns only weeks in which the teaching took place, be it in person or online. Generally, we were more strict in selecting the reliable covariate values as concerns Estimation procedures C and CC rather then D and DH.

**Table 3.**
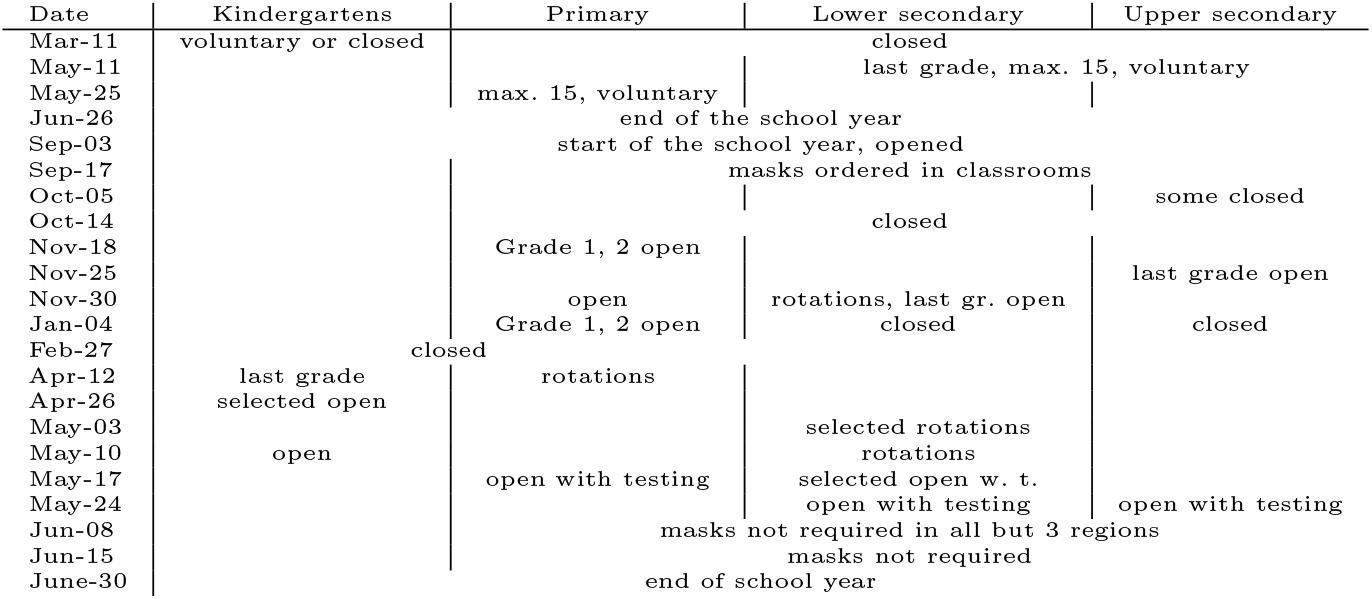
Schedule of the main mitigation measures at schools in 3/2020-6/2021.

Table 4 summarizes analyses we made for Comparisons C and CC. In Table 5 we list the covariate values for Comparison C and CC analyses, in Tables 6, 7 we present those for the D and DH analyses. Empty spaces in the Table 5 mean exclusion of the respective week. In the latter two tables, some values are determined for the sake of evaluating *ρ*_*t*_ in Figure 2, but are excluded from estimation, which fact is indicated by × symbol. One latter superscript symbols indicate reasons for exclusion or fractional values. Meaning of superscripts: a – voluntary, only 15 pupils in a classroom (approx. half), b – only 4 days from week, c – only 1st and 2nd classes open, d – only the last year open, e – summer vacation, f – Christmas vacation, g – autumn vacation, h – closed on Wednesday, i – masks ordered in classrooms, j – different regime among regions according to epidemiological situation, k – regime change during a week, l – last grade fully open, the rest on rotations, m – starting from Tuesday, valid in all but 3 regions on Monday, n – open up to decision of school principals.

**Table 4.** The comparison analyses. For kindergartens, 26/11–1/12 are omitted due to Autumn vacation. In kindergarten cohort comparison, the control cohort is 7-year-olds and the “overall” cohort, i.e. the one by which the regime coefficients are multiplied, is the kindergarten one; this can be done as 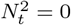 for all *t* used in the estimation.

|  | Kindergartens | Primary | Lower secondary | Upper secondary |
| --- | --- | --- | --- | --- |
| Group 1 | 5 years old | 7 years old | Grade 9 | Grade 13 |
| Sub-cohort 1 | $Z^5$ | $Z^7$ | $\frac{1}{2}(Z^{14} + Z^{15})$ | $\frac{1}{2}(Z^{19} + Z^{20})$ |
| Group 2 | 7 years old | 9 years old | Grade 7 | Grade 11 |
| Sub-cohort 2 | $Z^7$ | $Z^9$ | $\frac{1}{2}(Z^{12} + Z^{13})$ | $\frac{1}{2}(Z^{17} + Z^{18})$ |
| Period | 10/19 – 11/15/20 | 1/4 – 2/26/21 | 11/30 – 12/21/20 | 11/25 – 12/21/20 |
| Full weeks | 3 | 8 | 3 | 3 |
| C | | $\mu^1, \mu^2$ | $\mu^1, \rho^2$ | $\mu^1, \mu^2$ |
| CC | $\nu$ | $\mu$ | $\mu - \rho$ | $\mu$ |

**Table 5.**
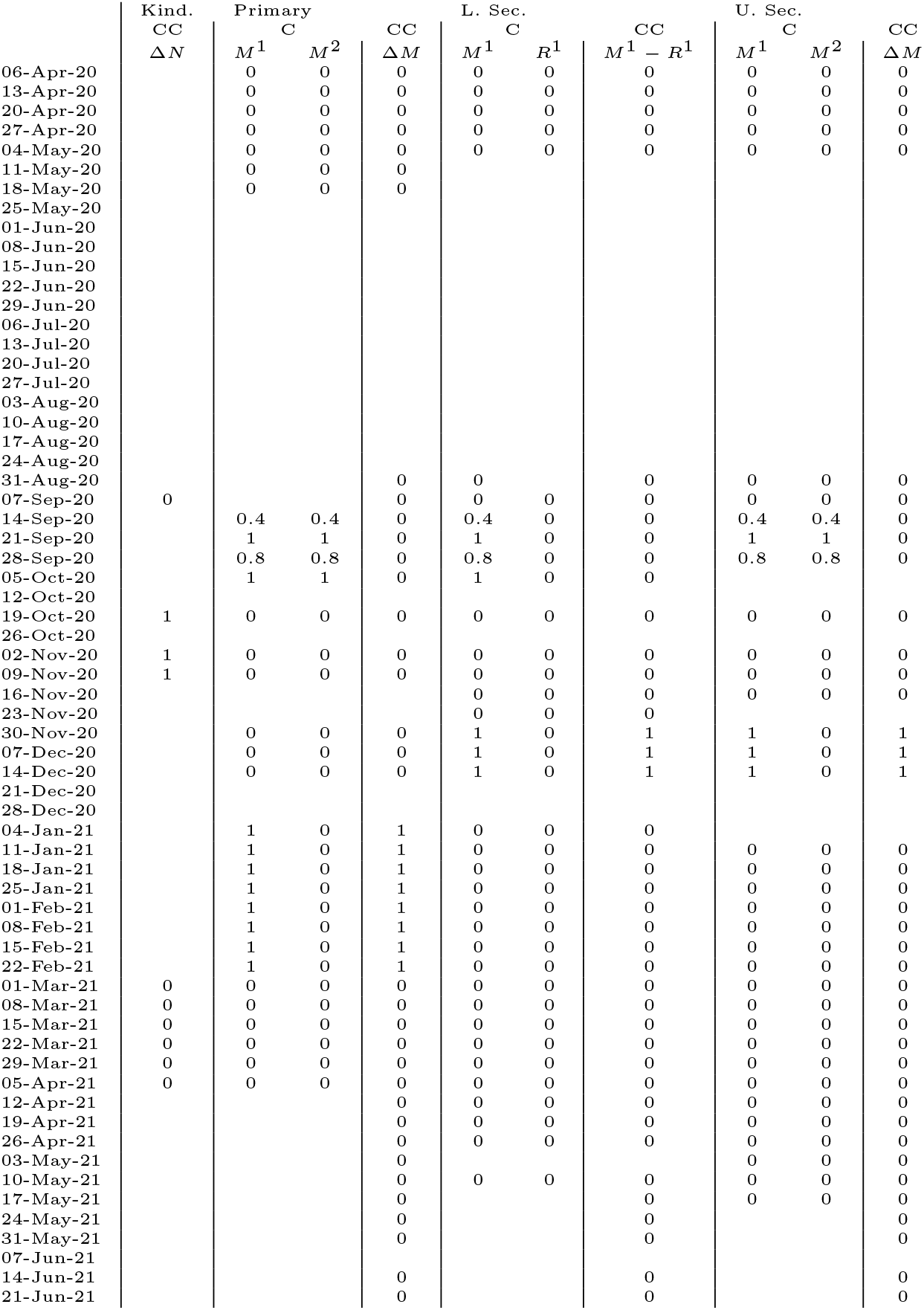
Covariates of the cohort analyses – Estimation procedures C (*M*^1^, *M*^2^, *R*^1^) and CC (Δ*M*, Δ*N, M*^1^ − *R*^1^).

**Table 6.**
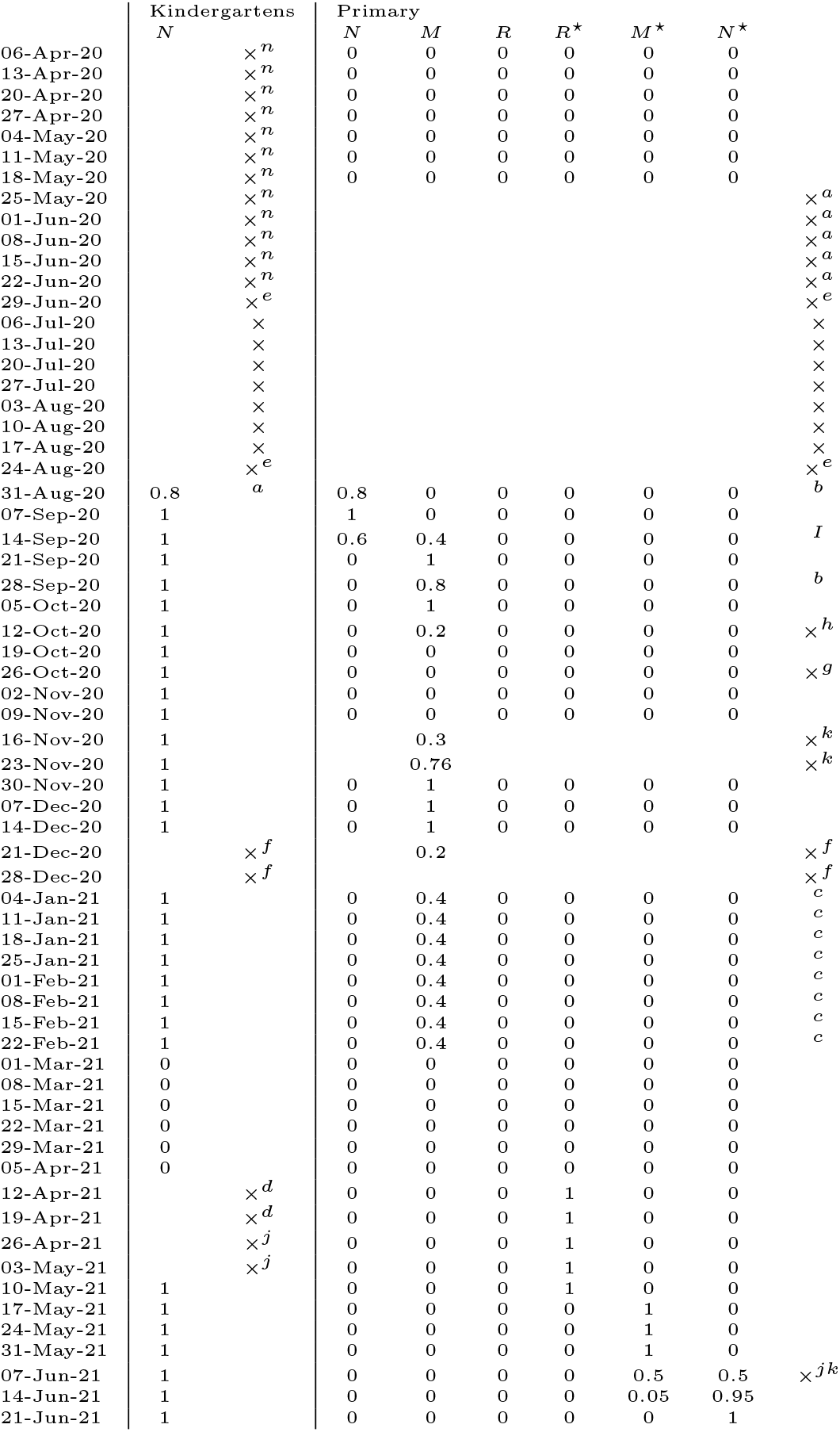
Covariates of D and DH – kindergartens and Grades 1-5.

**Table 7.**
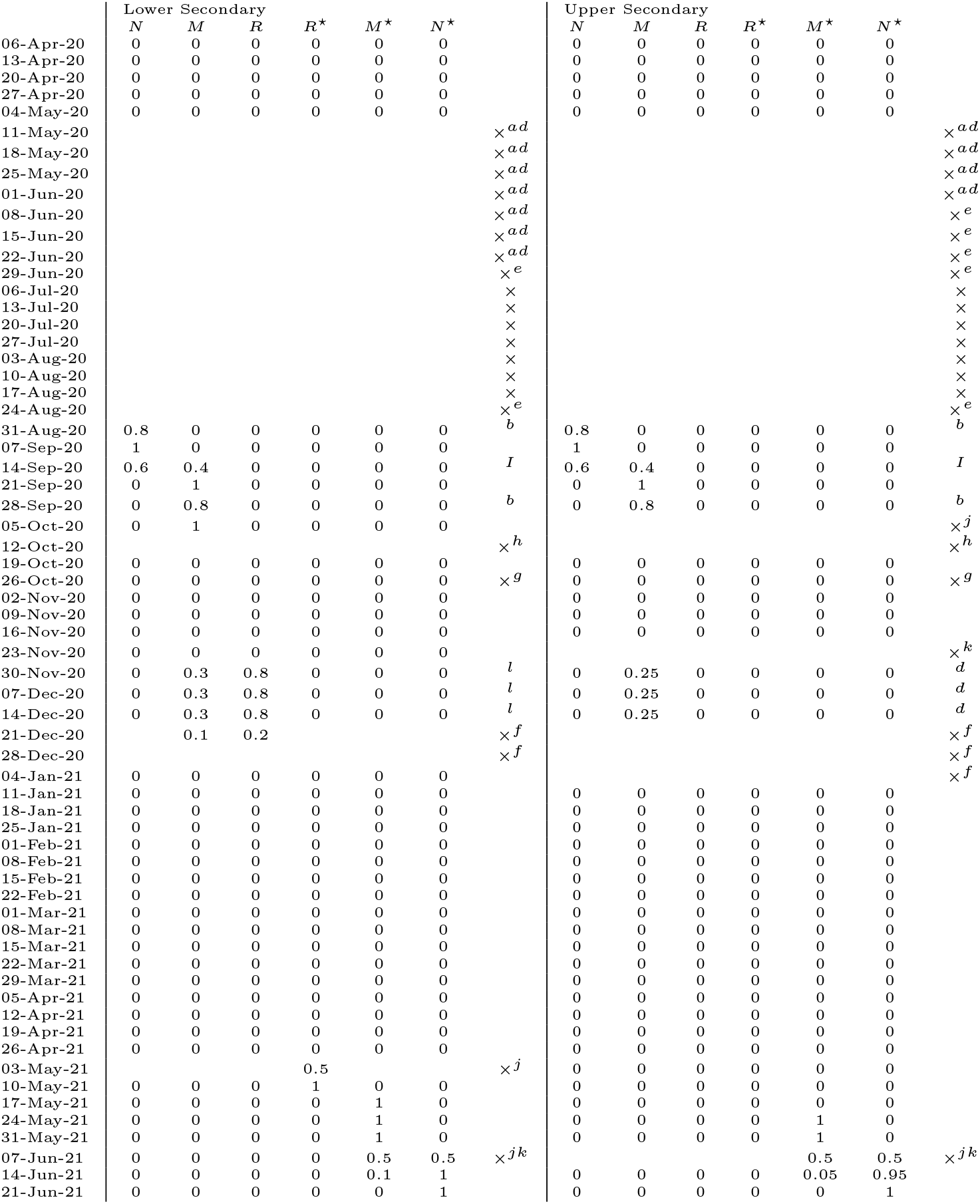
Covariates of D and DH – Grades 6 - 13.

### C Results

In all Estimation procedures C, CC, D and DH, we estimated coefficients by weighted least squares where we took *w*_*i,t*_ = max(20, *v*_*i*_*Y*_*t*−1_)^2^ as weights of residual variances (recall that *v*_*i*_ is the weight of the *i*-th district in the population). This choice was motivated by residual analysis and our desire to suppress observations suffering from the small-sample property.

The comparison of 5-year-olds and 7-year-olds, estimating *ν* at the kindergarten level, has been done in a slightly different way than it is described in Section A. The reason is that the sub-cohorts from different school levels are compared here. Generally, such comparison would not be correct as *X*^1^ depends on the cohort corresponding to kindergarten level (*X*^*kind*^) while *X*^2^ depends on the primary-level cohort (*X*^*prim*^). For this reason, Procedure C could not be applied. However, Procedure CC, in which we used only observations with *N*^2^ = 0 (primary schools closed, see Tables 5 and 6), could be done as we would have 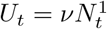 here, so we could have

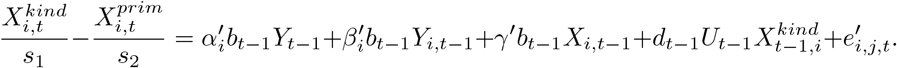

Table 8 shows the parameter estimates by individual models (recall that the regime coefficients always evaluate the rate of infections caused by the own cohort up to *d*_*t*_, so they are comparable each with the other coefficients as well as between the cohorts). We see in Table 8 that the vast majority of regime coefficients comes out positive and significant for all the types of schools. Figure 7 depicts these results graphically.

**Table 8.** Details of parameter estimation (SE in parenthesis)

| Model |  | Kindergartens | First degree | Second degree | Secondary |
| --- | --- | --- | --- | --- | --- |
| C<br>($w = 15\%$<br>Each) | $\nu^1$ | | | | |
| | $\nu^2$ | | | | |
| | $\mu^1$ | | 0.13*** (0.03) | 0.2*** (0.04) | 0.13** (0.05) |
| | $\mu^2$ | | 0.12** (0.06) | | 0.31*** (0.07) |
| | $\rho^2$ | | | 0.14** (0.06) | |
| CC<br>($w = 55\%$) | $\nu$ | 0.19*** (0.05) | | | |
| | $\mu$ | | 0.16*** (0.03) | | 0.16** (0.06) |
| | $\mu - \rho$ | | | 0.04 (0.06) | |
| D<br>($w = 10\%$) | $\nu$ | 0.22*** (0.02) | 0.15*** (0.05) | 0.56*** (0.04) | 0.89*** (0.05) |
| | $\mu$ | | 0.19*** (0.03) | 0.03 (0.04) | 0.27*** (0.05) |
| | $\rho$ | | | 0.05 (0.05) | |
| | $\rho^*$ | | 0.16*** (0.02) | 0.08* (0.04) | |
| | $\mu^*$ | | 0.14*** (0.03) | 0.17*** (0.03) | 0.47*** (0.04) |
| | $\nu^*$ | | 0.11** (0.05) | 0.28*** (0.04) | 0.6*** (0.05) |
| | $\gamma$ | -0.14*** (0.04) | -0.1*** (0.03) | -0.09** (0.04) | -0.31*** (0.05) |
| DH<br>($w = 5\%$) | $\nu$ | 0.19*** (0.01) | 0.17*** (0.04) | 0.52*** (0.03) | 0.77*** (0.02) |
| | $\mu$ | | 0.22*** (0.02) | 0.02 (0.03) | 0.12*** (0.03) |
| | $\rho$ | | | 0.04 (0.05) | |
| | $\rho^*$ | | 0.13*** (0.02) | 0.05 (0.04) | |
| | $\mu^*$ | | 0.12*** (0.02) | 0.16*** (0.02) | 0.29*** (0.02) |
| | $\nu^*$ | | 0.07* (0.04) | 0.24*** (0.03) | 0.45*** (0.03) |
| | $\alpha$ | 0.05* (0.03) | 0.11*** (0.02) | 0.13*** (0.03) | 0.34*** (0.05) |
| | $\beta$ | 0.44*** (0.03) | 0.56*** (0.02) | 0.75*** (0.03) | 0.61*** (0.04) |

**Fig. 7.**
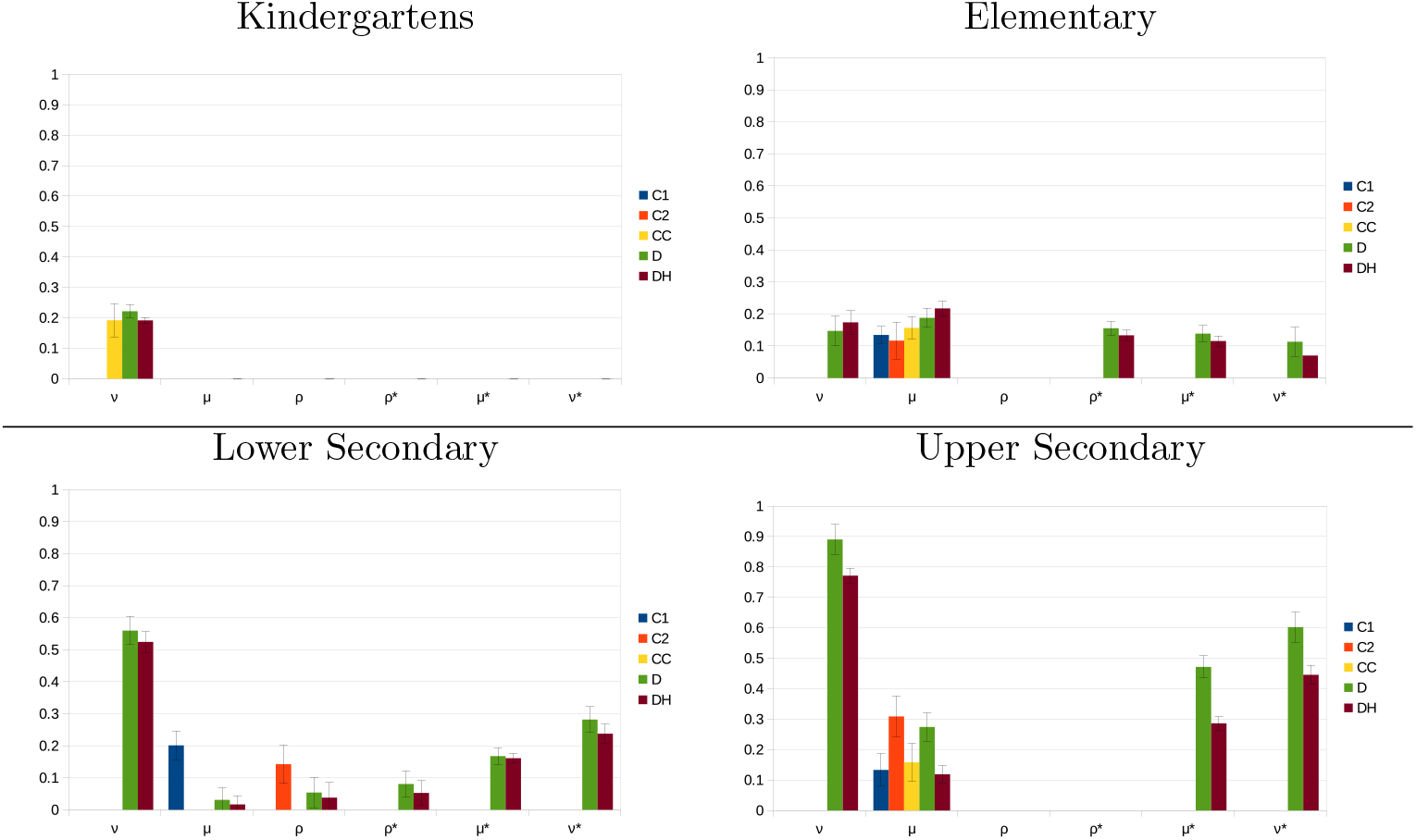
Regime coefficients

Having multiple estimates for the regime parameters from the four Estimation procedures C, CC, D and DH, we also computed meta-estimates of the regime parameters. These meta-estimates were computed as weighted averages of the estimates by the individual models, where we gave weight 55% to Procedure CC (which we regard as most independent of exogenous influences), 15% to each of the two estimates given by Procedure C, 10% to D and 5% to DH. The results may be found in Table 1 in the main text. As parameter *γ* (the influence of the same cohort independent of the school regime) came out mostly insignificant in the Procedure DH, we used version of DH without this parameter when computing the meta-estimates.

### D The Epidemic Model

To capture the influence of other infection sources than schools, we use a simple model in which the infections *X*_*i,t*_ in the cohort within the district *i* at time *t* depend on the previous week overall infections, the infections within the district and the infections within the cohort in the district:

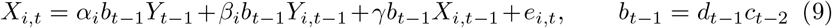

Here, *c*_*t*_ is the overall contact reduction reported by the longitudinal sociological study [18] and *d*_*t*_ is the rate of infectiousness. The time lag two weeks of the contact restriction was chosen as that maximizing the correlation: max_*τ*_ corr(*Y*_*t*_, *c*_*t*−*τ*_ *Y*_*t*−1_), the infection rate was determined as *d*_*t*_ = *w*_*t*_*r*_*t*_*i*_*t*_. Here, *w*_*t*_ = (1 + *ς* cos(*at* + *b*)) – where *ς* = 0.18, *a* and *b* are set so that the period is one-year period and the peak is on January, 10th – is a cyclic component reflecting the (direct or indirect) influence of weather conditions. Further, *r*_*t*_ is the course of infectiousness, determined by the composition of the virus variants present in Czechia, which we assumed to be constant, equal to *r*^0^ = 1.55, up to the end of 2020, linear up to March-01-2021 (due to alpha variant onset), and then constant, equal to *r*^1^ = 2.44. Finally, *i*_*t*_ = (1 − *αι*_*t*_) is the effect of natural immunization, where *α* = 0.4 is the ascertainment rate and *ι*_*t*_ is the ratio of total reported infections within the examined cohorts (i.e. children from age 4 to 19 and the half of the cohort of 20-year-olds). As no children younger than 16 were vaccinated by June 2021 and only a small minority of students over 16 had got their first dose by that time, we did not include the effect of vaccination in *d*_*t*_. The parameters *ς, r*^0^, *r*^1^ we obtained by estimation; notably, the value of *ς* is very close to that obtained independently by [24]. The ascertainment rate *α* has been set according the rate of respondents of [18] suffering from typical covid symptoms who underwent testing. See Figure 9 for the course of *d*_*t*_ and its empirical counterparts, and also Table 9 for the estimation of homogenized version of (9):

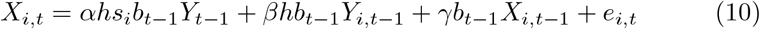

where *h* is the relative size of the examined cohort with respect to the rest of the population and *s*_*i*_ is the relative size of the *i*-th district’s population.

**Table 9.**
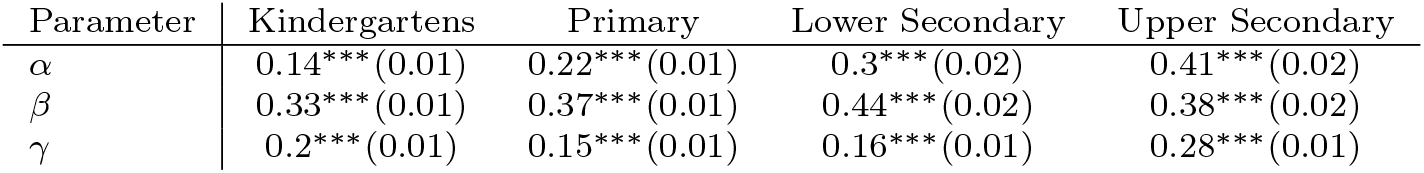
Estimation of (10)

| Parameter | Kindergartens | Primary | Lower Secondary | Upper Secondary |
| --- | --- | --- | --- | --- |
| $\alpha$ | 0.14*** (0.01) | 0.22*** (0.01) | 0.3*** (0.02) | 0.41*** (0.02) |
| $\beta$ | 0.33*** (0.01) | 0.37*** (0.01) | 0.44*** (0.02) | 0.38*** (0.02) |
| $\gamma$ | 0.2*** (0.01) | 0.15*** (0.01) | 0.16*** (0.01) | 0.28*** (0.01) |

**Fig. 8.**
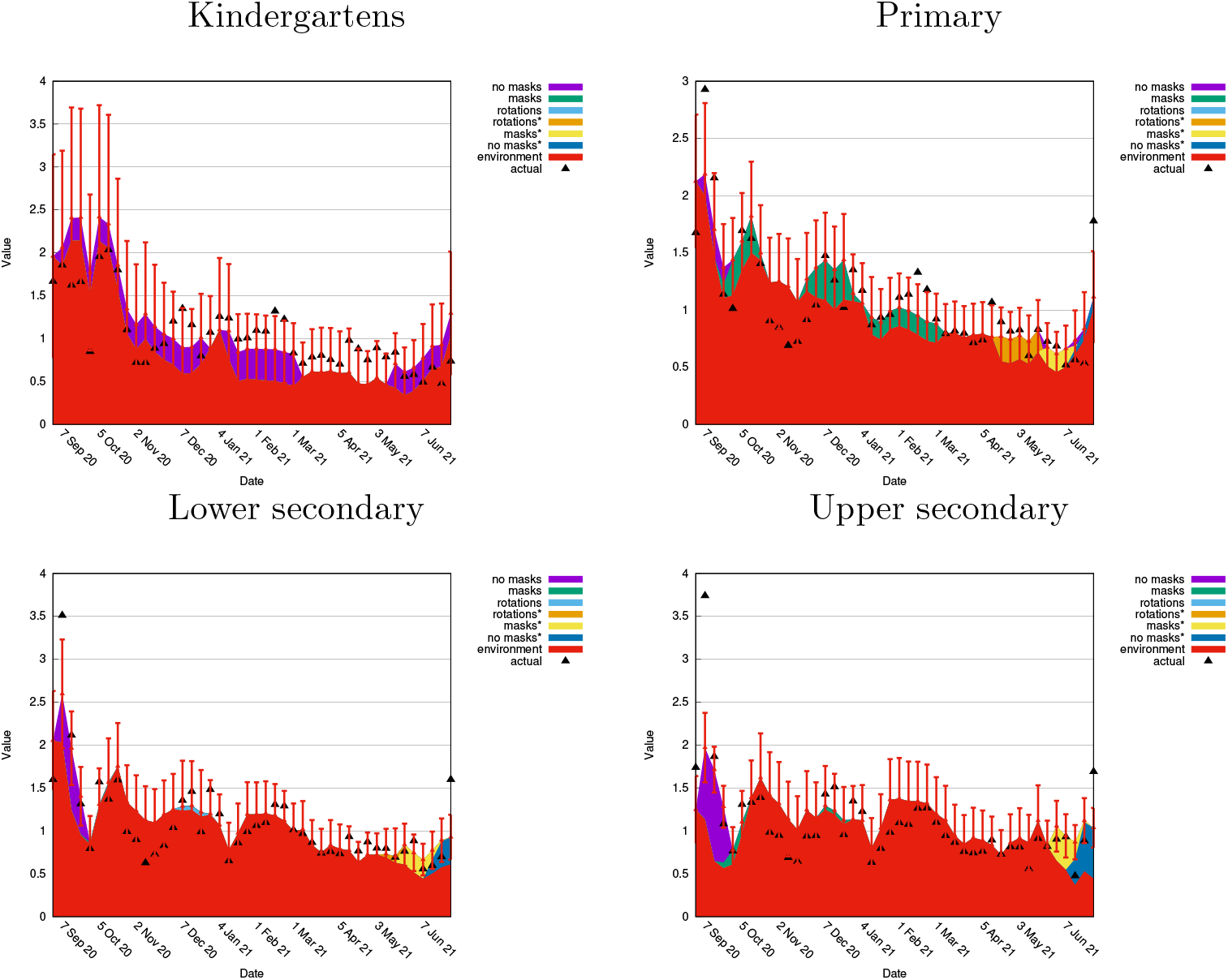
Growth 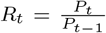, its one-week prediction *ρ*_*t*_ and its decomposition, based only on Procedure DH. Errorbars: 95% prediction CI

**Fig. 9.**
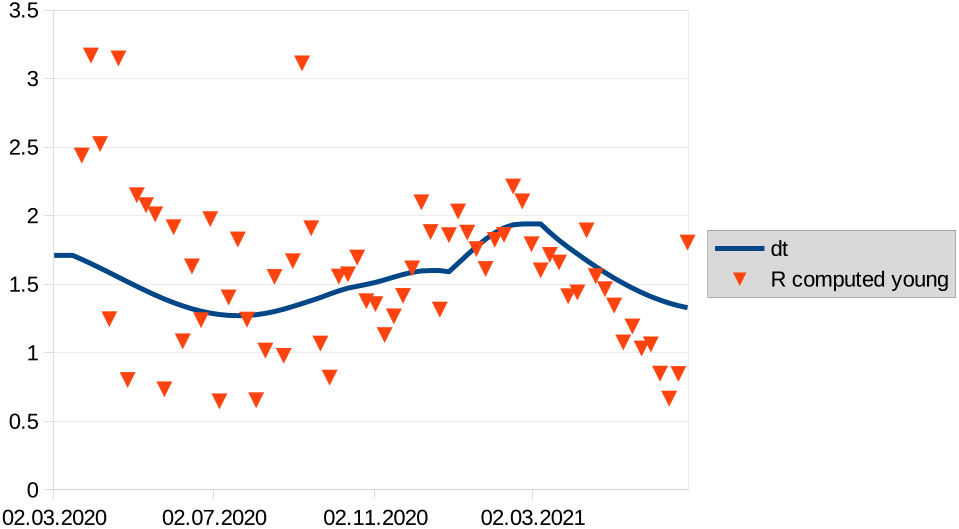
Assumed *d*_*t*_ (line) and empirical *d*_*t*_ (triangles). The empirical one is estimated by 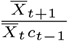,where 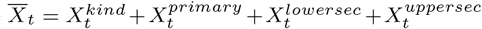 is the number of infections in all the examined cohorts. The sudden increase at the beginning of 2021 is due to the alpha variant.

### E Sensitivity analysis – variation of *d*_*t*_

A question may arise, to what extent our results are dependent on the choice of model parameters. In the present section, we show how the results change when the “course of infection” *d*_*t*_ is altered, namely when the “amplitude” parameter *ς*, originally equal to 0.18 (see Section D), is doubled, i.e. *ς* = 0.36. Figure 10 shows that the altered curve may be regarded as valid; it fits better the actual data in spring 2021 for the price of a worse fit in winter. Figure 11 shows the results given the altered *d*_*t*_. A comparison with Figure 1 shows that even though the actual values vary slightly, their ratio is similar.

**Fig. 10.**
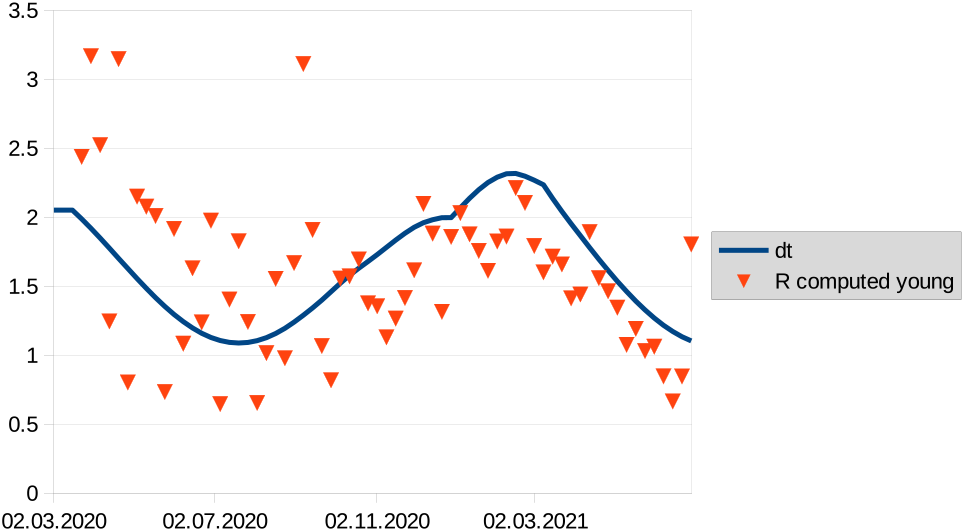
Altered *d*_*t*_. See Figure 9 for a legend.

**Fig. 11.**
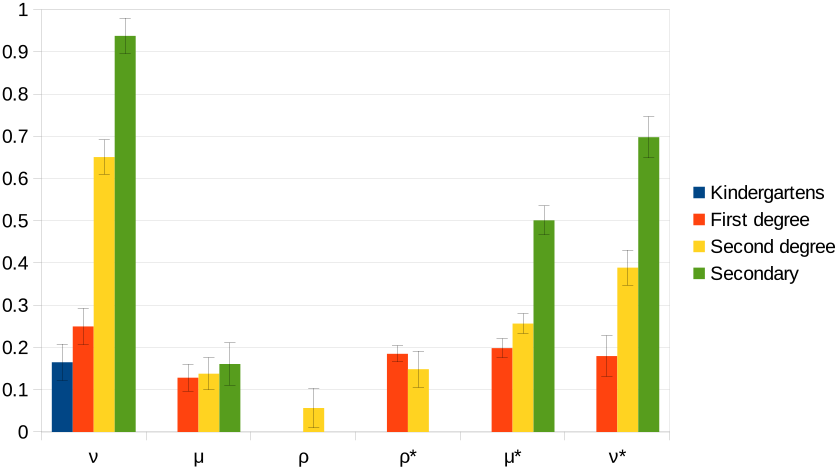
Meta-estimates of regime coefficients for altered *d*_*t*_ (*ς* = 0.36)

### F Ascertainment Rate

It is widely admitted that Covid-19 infections are reported only partially. In this section we show that our methods are robust with respect to this fact.

First we discuss the situation when the ascertainment rate is the same among cohorts and over time which means that there is a constant 0 *< α* ≤ 1 such that 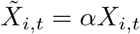 where 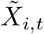 is the number of reported cases in district *I* at time *t*, and similarly with 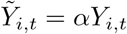 and 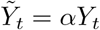. Then, by means of any Estimation procedure (C, CC, D, DH) we get the correct coefficients because (4) lacks a constant term, thanks to which the true values satisfy (4) if and only if the reported values satisfy this equation. Thus, if the ascertainment rate were the same for all cohorts over time, then using only reported values instead of the true ones for our estimation gives identical results.

Next we discuss the case in which the ascertainment rate is not the same for all cohorts and times. The smaller problem is when the rate fluctuates over time but remains the same over the cohorts, being equal to *α*_*t*_. Once we can assume that the fluctuation is random and the *α*_*t*_ is a martingale (i.e. the conditional expectation of *α*_*t*_ given the past up to *t* − 1 is *α*_*t*−1_), then using reported values instead of the true ones only adds a noise into the equation, but the estimation (by WLS) is still able to give correct results.^4^ If, alternatively, *α*_*t*_ were deterministic piece-wise constant with (unknown) jumps at (known) times *t* ∈ *T*, then (4) would hold for all *t ∉ T* ; thus it would suffice to omit the observations from times *T* to get correct estimates. Summarized, varying *α* does not prevent our methods to get correct estimates, yet it might require to drop some “suspicious” observations.

An additional complication could be that *α* differs among cohorts. Consider, for instance, the case when the rate is *α*^⋆^ within the examined school cohort and *α* within the whole population. Then, instead of (4), we would estimate

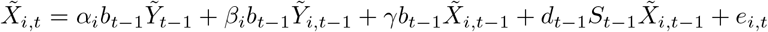

or, equivalently,

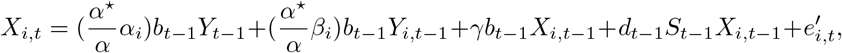

from which it is clear that the estimates of the regime coefficients would be correct; however, if we did not know the ratio 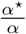, then the comparison of the estimated regime coefficients with with estimated *ω* (computed from *α*_*i*_ and *β*_*i*_) would not be correct.

Finally we discuss the case when the ascertainment rate grows with certain regimes. In particular, we assume that the overall ascertainment is *a*, but with introduction of certain regimes increases the ascertainment rate in the corresponding school cohort to *ϕa* where *ϕ >* 1 is a fixed constant. Denoting *I* the subset of times such that *t* ∈ *I* if and only if regimes increasing the ascertainment were fully introduced (i.e. with the corresponding covariates equal to one) both at *t* and *t* − 1, we get, for each *t* ∈ *I*,

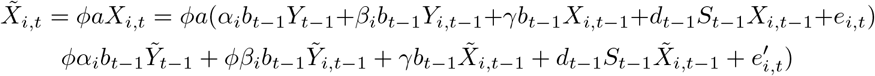

Further, for *t* ∈ *J*, where *J* is the set of all the times in which as well as one period back the regimes not increasing the ascertainment were fully introduced, the equation (4) holds for observed values instead of the true ones (see above). Putting this together, we get, for each *t* ∈ *I* ∪ *J*,

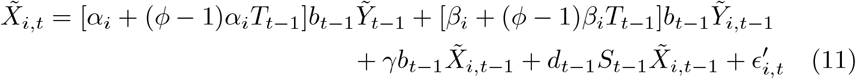

where *T*_*t*_ is an indicator of set *I* (i.e. it is equal to one if *t* ∈ *I* and is zero otherwise).

Using this model, we can tackle the hypothesis that opening of kindergartens increases the ascertainment rate in the kindergarten cohort (because of contact tracing and thus more testing in kindergartens). In particular, we can use a modified Procedure CC, which follows from (11):

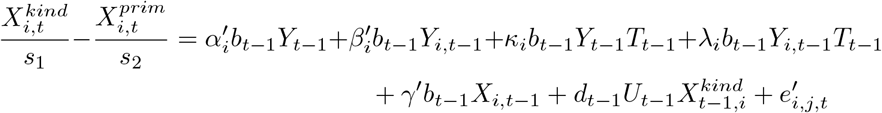

where *κ*_*i*_, *λ*_*i*_ is the additional set of coefficients. The estimate of *ν* obtained. this way is *ν* = 0.19, (*SE* = 0.16). This applies no matter the ascertainment rate and this is virtually the same value as in the main analysis; however, not significant this time, which is caused mainly by a large number of parameters of the modified Procedure (306 parameters per 693 observations). If we resort to the homogeneous analog of (11):

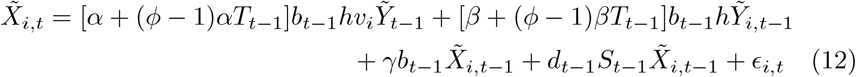

and use an analog of Procedure CC, we get a sharper value 0.22 (*SE* = 0.09), which is again in line with the main analysis.

Another ascertainment-related objection concerning our results could be that the ascertainment is naturally higher when the regular testing take places in schools (which is the case of regimes denoted by star in our notation). We explored this hypothesis using (12) for *t* ∈ *I* ∪ *J* where *I* and *J* are the times such that for *t* ∈ *I* and *t* ∈ *J* regular testing took place, did not take place, respectively, at both *t* and *t* − 1. The estimation has been done by Nonlinear Weighted Least Squares. Results of the estimation for all the levels but kindergartens can be seen in Table 10. The results indicate that, for primary and lower secondary level, regular testing is likely to increase ascertainment rate (i.e., *ϕ >* 1), and the effects of regimes with regular testing on true infections are likely to be lower than the effects on the reported numbers. For upper secondary schools, on the other hand, such effect has not been detected.

**Table 10.** Estimation of Procedure DH with the ascertainment rate multiplied by *ϕ* for the regimes with regular testing (we assumed *γ* = 0).

|  | Primary | Lower secondary | Upper secondary |
| --- | --- | --- | --- |
| $\nu$ | 0.18*** (0.04) | 0.53*** (0.03) | 0.77*** (0.02) |
| $\mu$ | 0.23*** (0.02) | 0.02 (0.03) | 0.12*** (0.03) |
| $\rho$ | | 0.04 (0.05) | |
| $\rho^*$ | 0.07*** (0.03) | -0.01 (0.05) | |
| $\mu^*$ | 0.08*** (0.02) | 0.12*** (0.02) | 0.32*** (0.03) |
| $\nu^*$ | 0.04 (0.04) | 0.21*** (0.03) | 0.45*** (0.04) |
| $\alpha$ | 0.1*** (0.02) | 0.12*** (0.03) | 0.36*** (0.05) |
| $\beta$ | 0.56*** (0.02) | 0.74*** (0.03) | 0.6*** (0.04) |
| $\phi$ | 1.16*** (0.08) | 1.17*** (0.08) | 0.96*** (0.12) |

Summarized, once we are interested in the true infections (i.e., including those unreported) rather than in the reported ones, our method can be used, too, as it is rather robust with respect to the ascertainment rate provided that it varies slowly and does not depend on the school opening regime. When the rate varies with the regimes, our Procedures D and DH, as well as C and CC in some special cases, may be modified to handle this issue.

## Footnotes

1 An alternative approach could be to compute *ρ*_*t*_ only by means of the estimates by Procedure DH; however, the results are similar, see Figure S8.

2 Note also that the results of the estimation can also be used to test the null hypothesis that the corresponding regime has no influence on infections, which will be rejected by significant values of either *µ*^1^ or *µ*^2^. However, the significance level has to be corrected here.

3 Again, the significant values may serve for the proof that the opening in the particular regime influences infections.

4 More detailed analysis would be necessary to study the properties of our estimates.

## References

[1] Auger, K.A., Shah, S.S., Richardson, T., Hartley, D., Hall, M., Warniment, A., Timmons, K., Bosse, D., Ferris, S.A., Brady, P.W., et al.: Association between statewide school closure and covid-19 incidence and mortality in the us. Jama 324(9), 859–870 (2020)

[2] Brauner, J.M., Mindermann, S., Sharma, M., Johnston, D., Salvatier, J., Gavenčiak, T., Stephenson, A.B., Leech, G., Altman, G., Mikulik, V., et al.: Inferring the effectiveness of government interventions against covid-19. Science 371(6531) (2021)

[3] Haug, N., Geyrhofer, L., Londei, A., Dervic, E., Desvars-Larrive, A., Loreto, V., Pinior, B., Thurner, S., Klimek, P.: Ranking the effectiveness of worldwide COVID-19 government interventions. Nature Human Behaviour 4(12), 1303–1312 (2020). https://doi.org/10.1038/s41562-020-01009-0. Publisher: Nature Publishing Group. Accessed 2021-09-25

[4] Bignardi, G., Dalmaijer, E.S., Anwyl-Irvine, A.L., Smith, T.A., Siugzdaite, R., Uh, S., Astle, D.E.: Longitudinal increases in childhood depression symptoms during the covid-19 lockdown. Archives of Disease in Childhood 106(8), 791–797 (2021)

[5] Di Pietro, G., Biagi, F., Costa, P., Karpiński, Z., Mazza, J.: The likely impact of covid-19 on education: Reflections based on the existing literature and recent international datasets. Technical report (2020). https://core.ac.uk/download/pdf/343468109.pdf Accessed 2021-09-25

[6] ECDC: Covid-19 in children and the role of school settings in transmission - second update. Technical report (2021). https://www.ecdc.europa.eu/en/publications-data/children-and-school-settings-covid-19-transmission

[7] Ravens-Sieberer, U., Kaman, A., Erhart, M., Devine, J., Hölling, H., Schlack, R., Löffler, C., Hurrelmann, K., Otto, C.: Quality of life and mental health in children and adolescents during the first year of the covid-19 pandemic in germany: Results of a two-wave nationally representative study. Preprint available at SSRN 3798710 (2021). Accessed 2021-09-25

[8] Lessler, J., Grabowski, M.K., Grantz, K.H., Badillo-Goicoechea, E., Metcalf, C.J.E., Lupton-Smith, C., Azman, A.S., Stuart, E.A.: Household covid-19 risk and in-person schooling. Science 372(6546), 1092–1097 (2021)

[9] Gettings, J., Czarnik, M., Morris, E., Haller, E., Thompson-Paul, A.M., Rasberry, C., Lanzieri, T.M., Smith-Grant, J., Aholou, T.M., Thomas, E., et al.: Mask use and ventilation improvements to reduce covid-19 incidence in elementary schools—georgia, november 16–december 11, 2020. Morbidity and Mortality Weekly Report 70(21), 779–784 (2021)

[10] Sharma, M., Mindermann, S., Rogers-Smith, C., Leech, G., Snodin, B., Ahuja, J., Sandbrink, J.B., Monrad, J.T., Altman, G., Dhaliwal, G., et al.: Understanding the effectiveness of government interventions in europe’s second wave of covid-19. medRxiv https://doi.org/10.1101/2021.03.25.21254330, 03 (2021)

[11] Madewell, Z.J., Yang, Y., Longini, I.M., Halloran, M.E., Dean, N.E.: Household transmission of sars-cov-2: a systematic review and meta-analysis. JAMA network open 3(12), 2031756–2031756 (2020)

[12] SAGE: TFC: Children and transmission, update paper, 17 December 2020. Paper prepared by the Children’s Task and Finish Group (TFC) for the Scientific Advisory Group for Emergencies (SAGE). (2020). https://www.gov.uk/government/publications/tfc-children-and-transmission-update-paper-17-december-2020 Accessed 2021-08-25

[13] Okarska-Napiera-la, M., Mańdziuk, J., Kuchar, E.: Sars-cov-2 cluster in nursery, poland. Emerging infectious diseases 27(1), 317 (2021)

[14] Ismail, S.A., Saliba, V., Lopez Bernal, J., Ramsay, M.E., Ladhani, S.N.: SARS-CoV-2 infection and transmission in educational settings: a prospective, cross-sectional analysis of infection clusters and outbreaks in England. The Lancet Infectious Diseases 21(3), 344–353 (2021). https://doi.org/10.1016/S1473-3099(20)30882-3

[15] Varma, J.K., Thamkittikasem, J., Whittemore, K., Alexander, M., Stephens, D.H., Arslanian, K., Bray, J., Long, T.G.: Covid-19 infections among students and staff in new york city public schools. Pediatrics 147(5) (2021)

[16] Zimmerman, K.O., Akinboyo, I.C., Brookhart, M.A., Boutzoukas, A.E., McGann, K.A., Smith, M.J., Maradiaga Panayotti, G., Armstrong, S.C., Bristow, H., Parker, D., Zadrozny, S., Weber, D.J., Benjamin, D.K.: Incidence and Secondary Transmission of SARS-CoV-2 Infections in Schools. Pediatrics 147(4), 2020048090 (2021). https://doi.org/10.1542/peds.2020-048090. Accessed 2021-09-25

[17] Komenda, M., Karolyi, M., Bulhart, V., Žofka, J., Brauner, T., Hak, J., Jarkovský, J., Mužík, J., Blaha, M., Kubát, J., Klimeš, D., Langhammer, P., Daňková, Š., Májek, O., Bartůňková, M., Dušek, L.: COVID-19: Overview of the current situation in the Czech Republic. Disease Update [online]. Prague: Ministry of Health, Czech Republic. https://onemocneni-aktualne.mzcr.cz/api/v2/covid-19/osoby.csv. Accessed on June-20-2021, ISSN 2694-9423 (2020)

[18] PAQ research: Život během pandemie. https://zivotbehempandemie.cz/. Sociologická studie, Accessed 2021-05-13 (2021)

[19] Leng, T., Hill, E.M., Holmes, A., Southall, E., Thompson, R.N., Tildesley, M.J., Keeling, M.J., Dyson, L.: Quantifying within-school sars-cov-2 transmission and the impact of lateral flow testing in secondary schools in england. medRxiv https://doi.org/10.1101/2021.07.09.21260271 (2021)

[20] Galmiche, S., Charmet, T., Schaeffer, L., Paireau, J., Grant, R., Chény, O., Von Platen, C., Maurizot, A., Blanc, C., Dinis, A., et al.: Exposures associated with sars-cov-2 infection in france: A nationwide online case-control study. The Lancet Regional Health-Europe 7, 100148 (2021)

[21] Buonsenso, D., Munblit, D., De Rose, C., Sinatti, D., Ricchiuto, A., Carfi, A., Valentini, P.: Preliminary evidence on long covid in children. Acta Pediatrica 110(7), 2208–2211 (2021)

[22] for National Statistics, O.: Prevalence of ongoing symptoms following coronavirus (covid-19) infection in the uk: 1 april 2021. Technical report (2021). https://www.ons.gov.uk/peoplepopulationandcommunity/healthandsocialcare/conditionsanddiseases/bulletins/prevalenceofongoingsymptomsfollowingcoronaviruscovid19infectionintheuk/5august2021

[23] Blankenburg, J., Wekenborg, M.K., Reichert, J., Kirsten, C., Kahre, E., Haag, L., Schumm, L., Czyborra, P., Berner, R., Armann, J.P.: Mental health of adolescents in the pandemic: Long-covid-19 or long-pandemic syndrome? Available at SSRN 3844826 (2021)

[24] Gavenčiak, T., Monrad, J.T., Leech, G., Sharma, M., Mindermann, S., Brauner, J.M., Bhatt, S., Kulveit, J.: Seasonal variation in sars-cov-2 transmission in temperate climates. medRxiv (2021)

